# Conversion of osteoporotic-like vertebral fracture severity score to osteoporosis T-score equivalent status: a framework study for Chinese older men

**DOI:** 10.1101/2022.11.09.22282109

**Authors:** Yì Xiáng J. Wáng, Jason C.S. Leung, Patti M. S. Lam, Timothy C.Y. Kwok

**Affiliations:** Department of Imaging and Interventional Radiology, Faculty of Medicine, The Chinese University of Hong Kong, Shatin, New Territories, Hong Kong SAR, China; JC Centre for Osteoporosis Care and Control, Faculty of Medicine, The Chinese University of Hong Kong, Shatin, New Territories, Hong Kong SAR, China; Department of Medicine and Therapeutics, Faculty of Medicine, The Chinese University of Hong Kong, Shatin, New Territories, Hong Kong SAR, China

**Keywords:** Osteoporosis, osteoporotic vertebral fracture, bone mineral density, T-score, spine

## Abstract

**Introduction:** To define what portion of older community men with what severity of radiographic osteoporotic-like vertebral fracture (OLVF) correspond to what low T-score status.

**Methods:** There were 755 community Chinese men (age: 76.4 ±6.7 years, range: 65-98 years) with thoracic and lumbar spine radiograph, and hip and lumbar spine bone mineral density measures. For each vertebra in a subject, a score of 0, -0.5, -1, -1.5, -2, -2.5, and -3 was assigned for no OLVF or OLVF of <20%, ≥20∼25%, ≥25%∼1/3, ≥1/3∼40%, ≥40%–2/3, and ≥2/3 vertebral height loss, respectively. OLVFss was defined as the summed score of vertebrae T4 to L5. OLVFss and T-scores were ranked from the smallest to the largest values

**Results:** OLVFss of -2, -2.5, -3, corresponded to a mean femoral neck T-score of -2.297 (range: -2.355∼-2.247), -2.494 (range: -2.637∼ -2.363), and -2.773 (range: -2.898∼-2.643), a mean hip T-score of-2.311 (range: -2.420∼-2.234), -2.572 (range: -2.708∼-2.432), -2.911 (range: -3.134∼-2.708), a mean lumbar spine T-score of -2.495 (range: -2.656∼-2.403), - 2.931 (range: -3.255∼-2.656), and -3.369 (range: 3.525∼-3.258). The Pearson correlation value of OLVFss and T-score of femoral neck, hip and lumbar spine was r =0.21, 0.26, and 0.22 (all p<0.0001).

**Conclusion:** A single severe grade OLVF (≥40% height loss) or OLVFss ≤-2.5 suggest osteoporosis suggest this subject is osteoporotic, or a single collapse grade (≥2/3 height loss) radiological OLVF or OLVF≤-3 meets osteoporosis diagnosis criterion. The results further highlight the difficulty of diagnosing osteoporotic vertebral fracture among Chinese older men.

---

While a radiological ‘severe’ osteoporotic vertebral fracture (OVF) would be conceptually considered a ‘clinically relevant’ fragility fracture, whether and how one or multiple ‘moderate’ radiological OVF together constitute a ‘clinically relevant’ fragility fracture has not been fully illustrated. Among Caucasian women, the lifetime risk of hip fracture at the age of 50 years is considered to be around 16%, while 30% is a conservative estimate of the risk of any osteoporotic fractures [1]. According to the WHO criteria, T-score is defined as: (BMD_patient_ – BMD_young normal mean_)/SD_young normal population_. In adult women, the cutpoint value of patient BMD 2.5 SD below BMD_young normal mean_ satisfies that, when the femoral neck is measured, osteoporosis prevalence is about 16.2% for those age ≥50 years, the same as the lifetime risk of hip fragility fracture [1]. If other sites are also considered, this cutpoint value identifies approximately 30% of postmenopausal women as having osteoporosis, which is approximately equivalent to the lifetime risk of fragility fracture at the spine, hip, or forearm. Following the same line of thinking as the WHO BMD definition, recently we performed a study to define what portion of older community women with what severity of radiographic osteoporotic-like vertebral fracture (OLVF) correspond to what low T-score status, i.e., we explore how we can convert severity of radiological OVFs to the equivalent T-score values [2]. Following our results for older women, in the current study we studied what portion of older community men with what severity of radiographic OLVF correspond to what low T-score status.

## Material and Method

From our MrOS(Hong Kong) baseline database of 2000 male subjects, we randomly selected 496 cases following the principle of one out of every five cases and also excluding the cases used in the year-14 follow-up study and those of poor radiograph quality. From the MrOS(Hong Kong) year-14 follow-up results, there were 259 male subjects. Putting these together, in total there were 755 male subjects, aged 76.4±6.7 years (range: 65-98 years).

On spine radiograph, vertebrae T4–L5 were evaluated with an extended version of semi-quantitative (eSQ) scheme with the following criteria [3]: (1) minimal grade refers to radiological osteoporotic-like vertebral fracture (OLVF) with < 20% height loss, which would be theoretically equivalent to SQ grade 0.5; (2) mild grade OVF is the same as Genant mild grade (≥20∼25% height loss); (3) Genant moderate grade OVF is divided into two subgrades: ≥25%∼1/3 height loss (moderate grade) and ≥1/3∼40% height loss (moderately-severe grade); (4) Genant severe grade OVF is divided into two subgrades: ≥40%∼2/3 height loss (severe grade) and with > 2/3 height loss (collapsed grade). Nonfractural changes of the vertebrae shape were systematically differentiated from OLVF as much as possible. How to grade OVF in men has not been clearly defined, though there has been sufficient evidence to suggest minimal and mild grade VDs in men are not associated with increased fracture risk statistically [4, 5]. We decided to count minimal and mild grade VDs initially, and then determine that limited number of minimal and mild grade VDs in a subject are not sufficiently clinically relevant.

An OLVF sum score (OLVFss), which describe the overall number and severity of osteoporotic fracture status of the thoracic and lumbar spine, was calculated for each study subject. For each vertebra in a subject, a score of 0, -0.5, -1, -1.5, -2, -2.5, and -3 was assigned for no OLVF or OLVF of <20%, ≥20∼25%, ≥25%∼1/3, ≥1/3∼40%, ≥40%–2/3, and ≥2/3 vertebral height loss, respectively. However, two adjacent minimal OLVFs were assigned as -0.5. Three adjacent minimal OLVFs are generally rare and was assumed to -1 [2]. OVFss was calculated by summing up the scores of vertebrae T4 to L5.

## Results

11.1%, 7.9%, 4.5% of the subjects had OVFss ≤ -2, ≤ -2.5, and ≤ -3 respectively.

OVFss of -2.0 corresponded a mean femoral neck T-score of -2.297 (range: -2.355∼-2.247), a mean total hip T-score of -2.311 (range: -2.420∼-2.234), and a mean lumbar spine T-score of 2.495 (range: -2.656∼-2.403).

OVFss of -2.5 corresponded to a mean femoral neck T-score of -2.494 (range: -2.637∼ - 2.363), a mean total hip T-score of -2.572 (range: -2.708∼-2.432), and a mean lumbar spine T-score of -2.931 (range: -3.255∼-2.656).

OVFss of -3 corresponded to a mean femoral neck T-score of -2.773 (range: -2.898∼-2.643), a mean total hip T-score of -2.911 (range: -3.134∼-2.708), and a mean lumbar spine T-score of -3.369 (range: 3.525∼-3.258).

The Pearson correlation value of OVFss and femoral neck T-score, total hip T-score and lumbar spine T-score was *r* =0.21 (95%CI: 0.14-0.28, p<0.0001), 0.26 (95%CI: 0.19-0.32, p<0.0001),and 0.22 (95%CI: 0.15-0.29, p<0.0001). respectively.

## Discussion

The results in this study suggest OLVFss ≤-2.5 suggests osteoporosis and OLVFss ≤-3 meets the criteria for osteoporosis diagnosis. Unsurprisingly, these results differ from our earlier results for women that OLVFss ≤ -1 suggests osteoporosis and OLVFss ≤ -1.5 meets the femoral neck T-score ≤ -2.5 criteria for osteoporosis diagnosis [3]. The high prevalence of vertebral deformity (VD) among healthy men has been well noted [5-7]. In order to provides ‘baseline noises’ VD information, recently we analyzed fracture shaped vertebral deformities (FSVDs) among 408 female patients and 374 male patients who had lateral chest radiographs due to mild illness or for routine healthcheck [23]. It was noted that FSVDs due to micro-fracture associated with physical stress are commonly seen among men. In that study, chest radiograph vertebral deformity index (VDI, including T4 to L1 or L2), which has the same principle in counting OLVFss, is shown in Fig-1. Fig-1 shows, VDI ≥-2.0 is very common among assumed normal BMD men, on the hand index ≤-3.0 tend to be uncommon. The current study suggests, even for a patient with a single OLVF of 40% height loss (VFss=-2.5), this patient may still not be necessarily osteoporotic. In our recent publication of MrOS (Hong Kong) year-4 follow-up, we demonstrated that, for elderly Chinese males (mean age 71.7 years, range 65–91 years), existing baseline OLVFs were only very weakly associated with higher risk of further OLVF [9]. When only the highest OLVF is counted at baseline, of subjects with Genant’s grade-0, 2.1% developed at least one OLVF progression or/and new OLVF, while of subjects with Genant’s grade-1, -2, and -3 OLVF, only 2.0% (3/149), 3.1% (3/96), and 2.8% (1/36) developed at least one OLVF progression/new OLVF, respectively. With a modified Genant SQ (semi-quantitative) method and OVF considered when a reduction in vertebral height and/ or compression of ≥10% of the estimated vertebral body height, Kherad *et al*. [10] reported that in men with one or several OVFs, there were no significant differences in the presence of back pain in any ages, nor there were differences in the presence of back pain regarding type or number of fractures. Based on vertebral morphometry applied to DXA derived images, Waterloo *et al*. [11] reported that presence of OVF in women was associated with an increased risk of back pain and lower quality of life score, but these associations were not present in men. In MrOS (USA) follow-up study (mean base-line age: 72.9 years), Ensrud *et al*. [12] reported 13.5% of incident SQ radiographic OVF in elderly men were also clinically diagnosed as incident fractures. In contrast, studies on elderly women suggest that one-quarter to one-third of incident radiographic OVF are also clinically diagnosed as fracture events. For example, in the postmenopausal women attending the Fracture Intervention Trial Research study, Fink *et al*. [13] reported that about 25% of incident radiographic OVF were also diagnosed as clinical VF. These findings may suggest VD in men might have been over-diagnosed as OVF. While OVF prevalence of Caucasian males is approximately half of that Caucasian females, OVF prevalence of Chinese males is further half of that of Caucasian males [14]. These factors further increase the difficulty of diagnosing true osteoporotic fracture of the spine among Chinese males.

**Fig-1.**
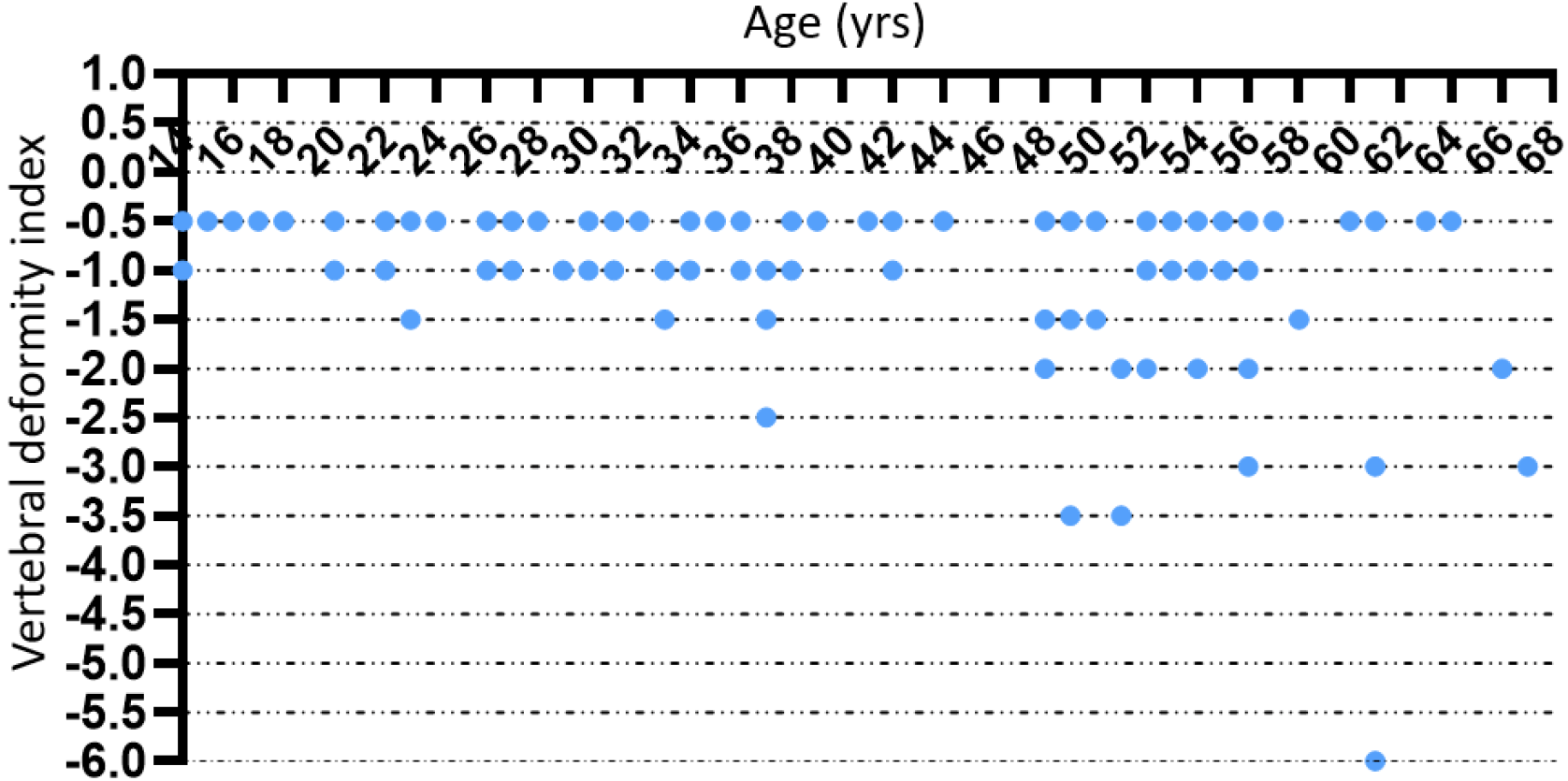
Chest radiograph vertebral deformity index (VDI) among young and middle-aged male patients (n=374) unrelated to spine disorders. It can be assumed that for men younger than 60 years old there would be few cases, or none, had osteoporosis. One vertebral deformity of ‘minimal’, ‘mild’, ‘moderate’, ‘moderately severe (mod/s)’, ‘severe’, ‘collapsed’ has <20%, ≥20∼25%, ≥25%∼1/3, ≥1/3∼40%, ≥40%∼2/3, and ≥2/3 vertebral height loss, and is assigned a score of 0.5, 1.0, 1.5, 2.0, 2.5, 3 respectively. Chest VDI is calculated by summing the scores of all vertebrae (T4 to L1 or L2). Patients with any fracture shaped vertebral deformity are presented.

In conclusion, for older Chinese men, a single severe grade OLVF (≥40% height loss) or OLVFss ≤-2.5 suggest osteoporosis suggests this subject is osteoporotic, or a single collapse grade (≥2/3 height loss) radiological OLVF or OLVFss ≤-3 meets osteoporosis diagnosis criteria. These results further highlight the difficulty of diagnosing OVF among Chinese older men.

## Data Availability

All data produced in the present study are available upon reasonable request to the authors

